# Racial and Ethnic Differences in the Determinants of Antibiotic Use for Acute Gastroenteritis in the United States

**DOI:** 10.64898/2026.02.04.26345585

**Authors:** Dongwook Kim, Raida Ismail, Finn Roberts, Gillian A.M. Tarr

## Abstract

**Background:** Most gastrointestinal infections do not require antimicrobial treatment, and the overall rate of use is low. However, differences in use across major American racial and ethnic groups is poorly understood. We estimated racial and ethnic disparities in antibiotic use for gastroenteritis and identified population-specific factors associated with use.

**Methods:** We analyzed the nationally representative 2018-2019 FoodNet Population Survey, limited to 1,950 individuals reporting gastroenteritis in the previous 7 days. Race and ethnicity were categorized as non-Hispanic Black, Hispanic, Other (Asian, Pacific Islander, American Indian and Alaska Native, and multiracial), and non-Hispanic White. Modified Poisson regression with survey weights estimated risk ratios for antibiotic use overall and within racial and ethnic groups.

**Results:** Antibiotic use was substantially higher among non-Hispanic Black (RR 4.6; 95% CI 4.01, 5.28), Hispanic (RR 2.2; 95% CI 1.00, 4.96), and Other (RR 3.8; 95%CI 2.27, 6.38) individuals, relative to non-Hispanic White respondents. Clinical factors were strongly associated with use in all racial and ethnic groups. However, socioeconomic associations qualitatively differed across groups; for example, higher income was associated with greater antibiotic use among Hispanic individuals but lower use among non-Hispanic Black individuals. Associations with social environment factors, such as Social Vulnerability Index and provider density, also varied by group.

**Conclusions:** We identified marked racial and ethnic disparities in antibiotic use, even after accounting for clinical and socioeconomic factors. Drivers of use differed by group, underscoring the need for tailored stewardship strategies and attention to the sociocultural factors influencing access to care and treatment decisions.

**Summary:** Antibiotic use for gastroenteritis is low; however, antibiotic use is significantly greater among individuals identifying as Black, Hispanic, or other races and ethnicities, relative to White individuals. Factors associated with use vary by population, emphasizing needed tailoring of stewardship strategies.

## BACKGROUND

Antimicrobial resistance (AMR) is a public health crisis, contributing to an estimated 4.95 million deaths in 2019 [1]. Multiple mechanisms are responsible for the rise of AMR, including the overuse of antibiotics in clinical settings. Overuse is particularly concerning in the context of acute gastroenteritis, where only a minority of cases will benefit from antimicrobial treatment [2–4]. Among individuals seeking care for gastroenteritis in primary care settings in the United States, 10-13% are treated with antibiotics, appearing to be of lower concern than overuse for conditions such as upper respiratory infections, where prescriptions can exceed 50% [5,6]. However, not all patients experience the same rate of antibiotic use. For gastroenteritis specifically, the only nationwide study of which we are aware to have examined patient characteristics found no racial or ethnic disparities in antibiotic prescribing, though it could not speak directly to antibiotic use [6]. For antibiotic use for other conditions, findings from prior studies focusing on racial and ethnic disparities are inconsistent, with two nationwide studies from similar periods showing opposite effects [7,8].

Here, we assess racial and ethnic disparities in antibiotic use for gastroenteritis, drawing on the Centers for Disease Control and Prevention (CDC) Foodborne Disease Active Surveillance Network (FoodNet) Population Survey. Importantly, we identify drivers of antibiotic use across and within different racial and ethnic groups, which can provide a basis for adapting antimicrobial stewardship programs and care practices to address health disparities [9].

## METHODS

### Study Design and Data Source

We conducted a retrospective cohort study using the CDC FoodNet Population Survey, conducted December 2017 through July 2019. At the time of the survey, FoodNet covered 15% of the U.S. population, including all of Connecticut, Georgia, Maryland, Minnesota, New Mexico, Oregon, and Tennessee, and select counties in California, Colorado, and New York [10]. The FoodNet Population Survey was conducted via random digit dialing telephone interviews to both land lines and cell phones and via a web-based survey.

Respondents provided sociodemographic information including their county of residence, zip code, socioeconomic characteristics, gastroenteritis signs and symptoms, care-seeking behavior, and potential exposures to pathogens monitored by FoodNet. Multiple versions of the survey were used, with respondents asked about their health in reference to either the previous 7 or 30 days. Due to greater anticipated accuracy of data relating to the previous 7 days, our primary analysis included only respondents who reported vomiting or diarrhea (gastroenteritis) in the previous 7 days. A secondary analysis examined respondents reporting gastroenteritis in the past 7 or 30 days.

We assigned the Social Vulnerability Index (SVI) (https://www.atsdr.cdc.gov/place-health/php/svi/svi-data-documentation-download.html) [11] and proportion of households reporting no access to a personal vehicle (https://www.census.gov/newsroom/press-kits/2019/acs-1year.html) based on each respondent’s county of residence. If county was missing, we assigned county based on zip code if a zip code was wholly included within a single county. As a proxy for access to care in an area, we calculated the rate of healthcare providers with a National Provider Identifier (NPI) per 1,000 population within a 10-km radius of the home zip code centroid.

This study was determined to not be human subjects research by the University of Minnesota IRB.

### Multiple Imputation

To address item nonresponse and avoid bias from listwise deletion, we used multiple imputation by chained equations [12]. The highest missingness was observed for bloody diarrhea (21.9%) and income (12.7%); vomiting, SVI, percent of households without a vehicle, and the NPI rate each had ∼7-8% missingness, and fever had 5.1%. Based on this level of missingness, we generated 25 imputed datasets using the mice package in R [12], specifying predictive mean matching for continuous variables, logistic regression for binary variables, and classification and regression trees for nominal variables with >2 categories.Convergence and plausibility diagnostics showed stable distributions across the 25 imputations.

### Statistical Analysis

Our primary outcome was self-reported antibiotic use for the identified gastrointestinal illness, potentially including both prescribed and unprescribed antibiotics. For the primary analysis, we modeled antibiotic use by demographic, socioeconomic, clinical, and social environment characteristics. Race and ethnicity were included in an overall model. Stratified models were built to estimate specific risk factors for antibiotic use among populations identified as non-Hispanic Black, Hispanic, another race or ethnicity, and non-Hispanic White, the categories FoodNet used to generate survey weights [10]. We estimated risk ratios (RRs) for antibiotic use with modified Poisson regression using a log link. We applied survey weights, which were generated to represent the target population of the FoodNet Population Survey accounting for nonresponse [10]. To account for correlation by interview mode (land line, cell phone, or web), we computed cluster-robust (Huber-White) standard errors. Each model was estimated for each multiply imputed dataset, and coefficients and cluster-robust variances were pooled using Rubin’s rules to obtain final estimates of coefficients, cluster-robust standard errors, and 95% confidence intervals (CIs). All analyses used R [13].

## RESULTS

The FoodNet Population Survey obtained completed surveys from 38,743 individuals. Of these, 1,950 (5.0%) reported vomiting and/or diarrhea in the previous 7 days; 3,346 (8.6%) additional respondents reported gastroenteritis in the previous 30 days. Among those reporting gastroenteritis in the previous 7 days, 92 respondents (4.7%) identified as non-Hispanic Black (“Black”), 156 (8.0%) as Hispanic, 1,534 (78.7%) as non-Hispanic White (“White”), 168 (8.6%) as another race or ethnicity (“Other”), and 18 (0.9%) did not report a race or ethnicity. Those in the Other racial and ethnic group primarily included participants identifying as Asian, American Indian or Alaska Native, or multiple races and/or ethnicities (Supplementary Table 1). All respondents with available data who reported antibiotic use (117/117) also reported seeking care for their gastroenteritis.

### Racial and Ethnic Disparities in Antibiotic Use

We observed pronounced racial and ethnic disparities in antibiotic use for gastroenteritis. Only 6.9% of White respondents reported antibiotic use for their illness, compared to 12.2% of Black, 14.3% of Hispanic, and 11.3% of Other respondents, not incorporating population weights (Table 1). From the weighted regression analysis, 5.3% of White, 24.2% of Black, 11.7% of Hispanic, and 20.0% of Other individuals with reference values for all other characteristics were estimated to use antibiotics. This equates to an adjusted 4.6-fold greater risk of antibiotic use among Black individuals (95% CI 4.01, 5.28), 2.2-fold greater risk among Hispanic individuals (95% CI 1.00, 4.96), and 3.8-fold greater risk among those of another race and/or ethnicity (95% CI 2.27, 6.38) (Figure 1).

**Table 1.** Characteristics of participants who reported experiencing gastroenteritis in the previous 7 days, stratified by racial and ethnic group.

|  | **Black,**<br>**Non-Hispanic**<br>**(n=90)** | **Hispanic**<br>**(n=154)** | **Other**<br>**(n=168)** | **White,**<br>**Non-Hispanic**<br>**(n=1,520)** |
| --- | --- | --- | --- | --- |
| **Antibiotic use, n (%)** |  |  |  |  |
| **No** | 79 (87.8) | 132 (85.7) | 149 (88.7) | 1,415 (93.1) |
| **Yes** | 11 (12.2) | 22 (14.3) | 19 (11.3) | 105 (6.9) |
| **Age, n (%)** |  |  |  |  |
| **Child <5** | 6 (6.5) | 15 (9.6) | 16 (9.5) | 59 (3.8) |
| **Child 5-17** | 6 (6.5) | 24 (15.4) | 36 (21.4) | 112 (7.3) |
| **Adult** | 80 (87.0) | 117 (75.0) | 116 (69.0) | 1,363 (88.9) |
| **Gender, n (%)** |  |  |  |  |
| **Man** | 25 (28.1) | 64 (43.2) | 69 (42.3) | 623 (41.3) |
| **Woman** | 64 (71.9) | 82 (55.4) | 93 (57.1) | 879 (58.3) |
| **Non-binary** | 0 (0.0) | 2 (1.4) | 1 (0.6) | 7 (0.5) |
| **Education, n (%)** |  |  |  |  |
| **High school** | 21 (22.8) | 42 (26.9) | 29 (17.3) | 268 (17.5) |
| **Some college** | 29 (31.5) | 44 (28.2) | 45 (26.8) | 429 (28.0) |
| **College graduate** | 42 (45.7) | 70 (44.9) | 94 (56.0) | 837 (54.6) |
| **Income, n (%)** |  |  |  |  |
| **≤\$39,999** | 43 (54.4) | 52 (37.1) | 43 (28.3) | 392 (29.5) |
| **\$40,000-74,999** | 15 (19.0) | 33 (23.6) | 35 (23.0) | 340 (25.5) |
| **≥\$75,000** | 21 (26.6) | 55 (39.3) | 74 (48.7) | 599 (45.0) |
| **Insurance, n (%)** |  |  |  |  |
| **No** | 10 (11.0) | 12 (7.9) | 8 (4.8) | 59 (3.9) |
| **Yes** | 81 (89.0) | 140 (92.1) | 159 (95.2) | 1,458 (96.1) |
| **Fever, n (%)** |  |  |  |  |
| **No** | 77 (89.5) | 122 (81.3) | 126 (78.8) | 1,289 (88.7) |
| **Yes** | 9 (10.5) | 28 (18.7) | 34 (21.2) | 165 (11.3) |
| **Vomiting, n (%)** |  |  |  |  |
| **No** | 58 (69.0) | 82 (59.4) | 102 (68.9) | 1,142 (80.3) |
| **Yes** | 26 (31.0) | 56 (40.6) | 46 (31.1) | 281 (19.7) |
| **Bloody diarrhea, n (%)** |  |  |  |  |
| **No** | 56 (86.2) | 91 (91.9) | 113 (95.0) | 1,196 (96.5) |
| **Yes** | 9 (13.8) | 8 (8.1) | 6 (5.0) | 43 (3.5) |
| **Immunocompromised,**<br>**n (%)** |  |  |  |  |
| **No** | 79 (85.9) | 137 (87.8) | 146 (86.9) | 1,367 (89.1) |
| **Yes** | 13 (14.1) | 19 (12.2) | 22 (13.1) | 167 (10.9) |
| **SVI, mean (SD)** | 0.55 (0.24) | 0.56 (0.24) | 0.46 (0.22) | 0.43 (0.23) |
| **Providers within 10 km radius (per 1,000 population), mean (SD)** | 9.53 (9.90) | 7.04 (5.67) | 8.10 (7.04) | 8.41 (9.63) |
| **Proportion of county population with no vehicle, mean (SD)** | 0.05 (0.04) | 0.03 (0.04) | 0.05 (0.06) | 0.03 (0.03) |

Abbreviations: SD, standard deviation

**Figure 1.**
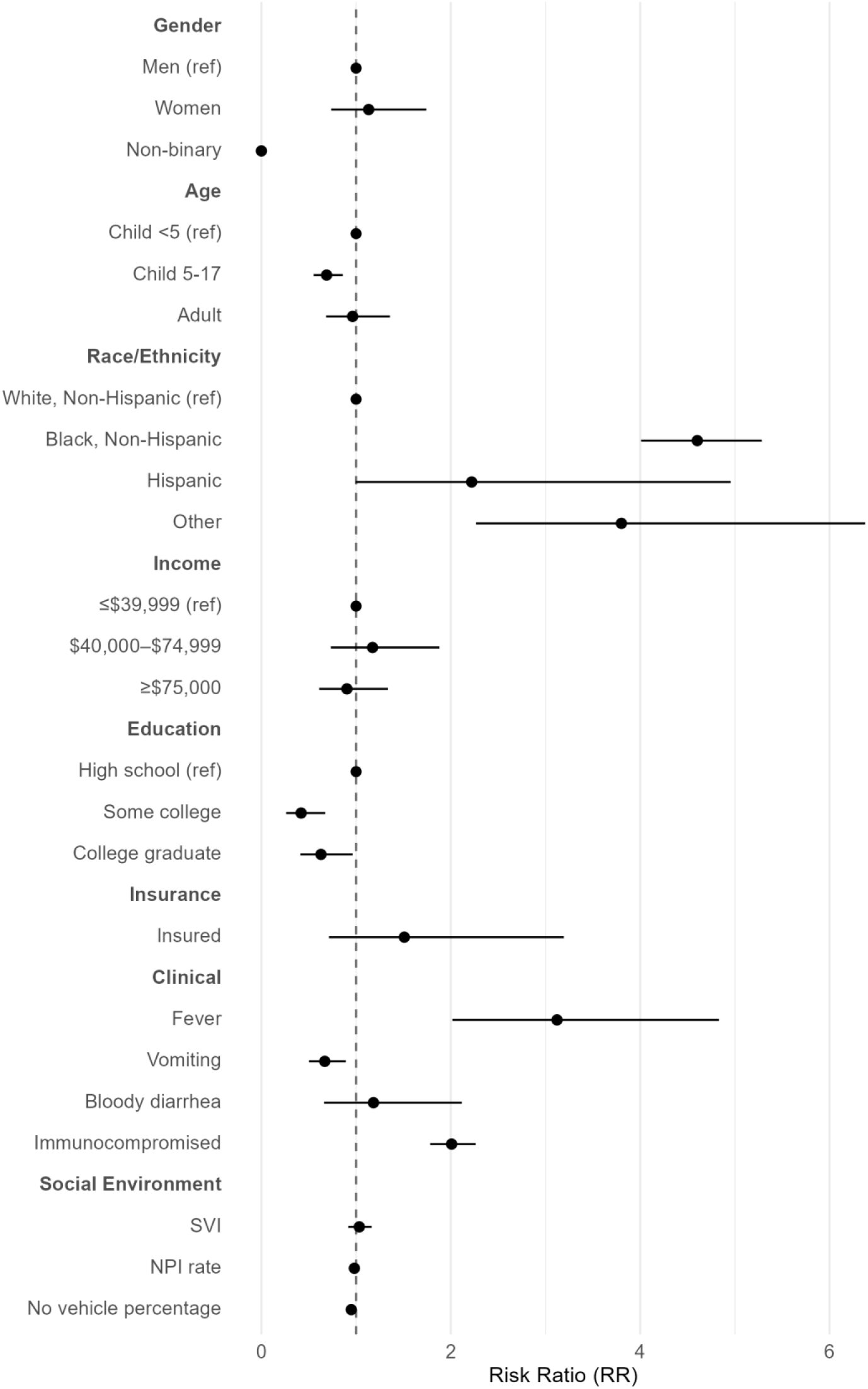
Risk ratios for antibiotic use of demographic, socioeconomic, clinical, and social environment characteristics. Risk of use was elevated for individuals identifying as non-Hispanic Black, Hispanic, or another race or ethnicity, relative to non-Hispanic White individuals. Dots indicate risk ratio (RR) point estimates and bars are 95% confidence intervals accounting for survey weights. For variables with >2 categories, the reference category is indicated, and for binary variables, the reference group is the absence of the characteristic. The RR for Social Vulnerability Index (SVI) is presented as the relative change in the proportion of individuals using antibiotics for each additional 0.1 increment in the index. The number of providers with a National Provider Identifier (NPI) per 1,000 population within a 10-km radius of the home zip code is presented as the relative change for each additional 1 provider. The percentage of a county with no personal vehicle is presented as the relative change for each additional 1%. **ALT Text**: Forest plot of risk ratio (RR) for antibiotic use. X-axis: 0-6. Higher risks for non-Hispanic Black (4.6) and Other (3.8) groups vs White reference. Fever and immunocompromise show notably high RRs, while income and social indicators stay near the reference line.

### Demographic Factors Had Minimal Impact on Antibiotic Use

Overall, children aged 5-17 years were significantly less likely to receive antibiotics than children <5 years (RR 0.69; 95% CI 0.55, 0.86), suggesting narrower prescribing among older pediatric patients with otherwise similar characteristics (Figure 1). This relationship was echoed in the Black and Other racial and ethnic groups (Figure 2). Conversely, only among the White population was age ≥18 years associated with lower antibiotic use, relative to children <5 years (RR 0.54; 95% CI 0.35, 0.83). Women were no more likely than men to take antibiotics for their gastroenteritis, and none of the individuals identifying as non-binary took antibiotics.

**Figure 2.**
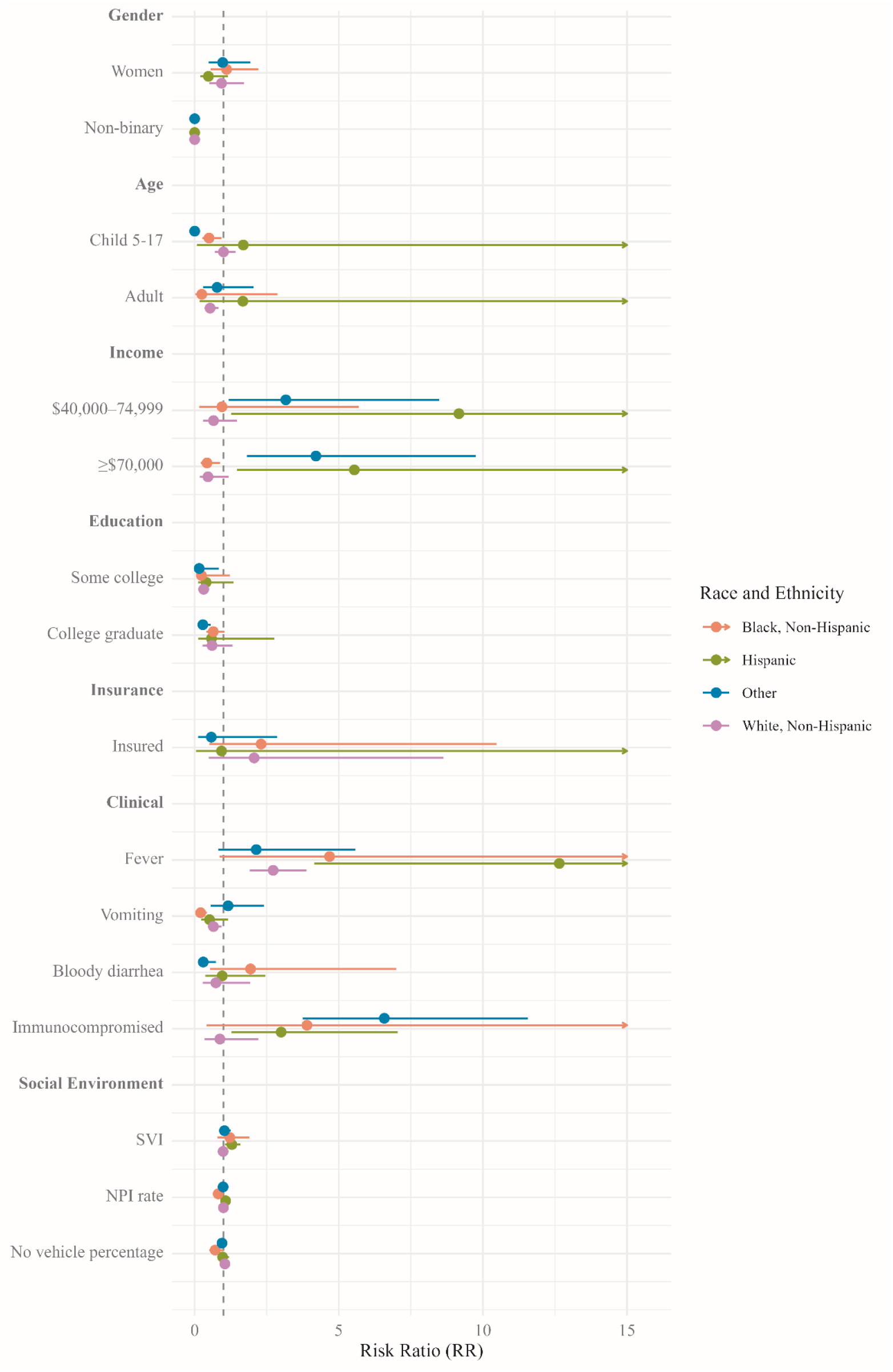
Risk ratios for antibiotic use of demographic, socioeconomic, clinical, and social environment characteristics, stratified by race and ethnicity. Drives of antibiotic use for gastroenteritis varied by race and ethnicity. Dots indicate risk ratio (RR) point estimates and bars are 95% confidence intervals (CIs) accounting for survey weights. Arrows on 95% CIs indicate that the right-hand side of the CI exceeds 15. For variables with >2 categories, reference categories are shown in Figure 1, and for binary variables, the reference group is the absence of the characteristic. RRs for Social Vulnerability Index (SVI) are presented as the relative change in the proportion of individuals using antibiotics for each additional 0.1 increment in the index. The number of providers with a National Provider Identifier (NPI) per 1,000 population within a 10-km radius of the home zip code is presented as the relative change for each additional 1 provider. The percentage of a county with no personal vehicle is presented as the relative change for each additional 1%. **ALT Text**: Stratified forest plot of antibiotic use risk ratio (RR) for non-Hispanic Black (orange), Hispanic (green), Other (blue), and White (purple). X-axis: -15; reference 1.0. Shows varied drivers by race/ethnicity. CIs for non-Hispanic Black and Hispanic groups exceeding 15 are marked with arrows.

### Importance of Socioeconomic Factors was Mixed

In combined analysis, educational attainment was associated with less antibiotic use among respondents who had some college (RR = 0.42; 95% CI 0.26, 0.67) or were college graduates (RR = 0.63; 95% CI 0.41, 0.96), relative to respondents with a high school degree, GED, or less (Figure 1). Risk ratios were similar across racial and ethnic groups, though were only statistically significant in the White and Other groups (Figure 2). We observed large associations between greater income and greater antibiotic use among Hispanic and Other respondents, with risk ratios ranging from 3.16 to 9.17 relative to individuals with incomes <$40,000 per year (Figure 2). However, this association was qualitatively different for Black individuals, with a RR of 0.43 (95% CI 0.21, 0.88) for individuals making ≥$75,000 per year. Estimates for the White population were similar but non-significant. Health insurance status was not significantly associated with antibiotic use.

### Clinical Characteristics Were Strongly Associated with Antibiotic Use

Appropriately, clinical features were among the strongest and most consistent risk factors for antibiotic use. Fever was associated with more than a threefold increase in the risk of antibiotic use (RR = 3.12; 95% CI 2.02, 4.83), and having an immunocompromising condition (e.g., HIV/AIDS, post-transplant status) doubled the risk (RR = 2.01; 95% CI 1.78, 2.26) (Figure 1). Conversely, vomiting was linked to lower use (RR = 0.67; 95% CI 0.50, 0.89). The strength and significance of these associations varied by racial and ethnic subgroup (Figure 2). For example, the Hispanic population had a 12.65-times greater risk of antibiotic use in the presence of fever (95% CI 4.15, 38.58), and having an immunocompromising condition increased risk by 6.58-fold (95% CI 3.75, 11.57) among those of other racial and ethnic groups. It was also only among this group that bloody diarrhea was associated with antibiotic use, with a 70% lower risk (RR 0.30; 95% CI 0.12, 0.74).

### The Social Environment Differed in Its Association with Antibiotic Use

Community context was associated with antibiotic use differently based on racial and ethnic group. The Hispanic population had a 1.29-fold increase in risk per 0.1-unit increase in SVI (95% CI 1.05, 1.59), but no other group experienced a difference in risk associated with SVI. Overall, there were modest but statistically significant decreases in risk associated with increases in NPI rate (RR = 0.98 per unit; 95% CI 0.97, 0.99) and the percentage of households in a county without a vehicle (RR = 0.95; 95% CI 0.92, 0.98). However, the Black population appeared to be more greatly affected by these factors, with an 18% decrease in antibiotic use per additional NPI provider per 10,000 population within a 10-km radius (RR 0.82; 95% CI 0.70, 0.95) and a 29% decrease per 1% increase in no-vehicle percentage (RR 0.71; 95% CI 0.51, 1.00). The association with no-vehicle percentage was qualitatively different for the White population, with a 5% increase in antibiotic use per 1% increase (RR 1.05; 95% CI 1.01, 1.08).

### Risk Factors for Gastroenteritis in the Previous 30 Days Were Similar

While models including respondents asked about their health in the previous 30 days had greater sample sizes (Supplementary Table 2), we considered the reliability of the longer recall period to be lower. Results of the secondary analysis were largely similar to those of the primary analysis, including elevated risk of antibiotic use among Hispanic, Black, and other racial and ethnic groups, relative to White respondents (Supplementary Figure 1). Notable differences from the primary analysis included increased risk of antibiotic use associated with having private insurance for Black and White respondents and decreased risk associated with vomiting for White and Other respondents (Supplementary Figure 2).

## DISCUSSION

We identified significant elevations in antibiotic use for gastroenteritis among Black (4.60) and other racial and ethnic groups (3.80), as well as a substantial but non-significant increase among Hispanic individuals (2.22), relative to the White group. This pattern places individuals in these racial and ethnic groups at greater risk of AMR, antibiotic-associated toxicity, and *Clostridioides difficile* infections. In addition to race and ethnicity, we also identified important overall differences in antibiotic use by age, level of education, clinical characteristics, proximity to prescribing providers, and community vehicle availability.

The magnitude of the racial and ethnic disparity in antibiotic use is greater than recognized in non-population-based studies. Nationwide [6] and single-site [14] studies found mixed or no differences between racial and ethnic groups in antibiotic prescribing. Although a substantial disparity has not previously been identified in gastroenteritis treatment, some studies of antimicrobial stewardship for other indications show greater prescribing in Black [15,16], Hispanic [15,16], and American Indian or Alaska Native [17] populations. Our results align more closely with these studies, though other studies show White populations with high antibiotic use [17–20]. The lack of consistency across previous studies suggests that context matters (condition, population, level of care). The disparities we identified persisted after adjustment, suggesting that measured socioeconomic, clinical and neighborhood factors explain only part of the observed differences. Remaining differences may stem from residual confounding in measured variables, variation in care seeking behaviors, the types of care settings accessed [21], and clinician characteristics [22].

Health literacy is an important component of appropriate antibiotic use [23,24], particularly in marginalized communities [25]. Both education and income are highly predictive of healthy literacy, with education having been identified as its greatest determinant [26,27]. However, higher education has also been associated with greater antibiotic use in high-income countries [28], and previous work has found no global association between income and antibiotic use [29]. This seemingly complex relationship between socioeconomic factors and antibiotic use is reflected in our results. Consistent with the hypothesis that higher socioeconomic status improves health literacy, Black respondents with higher incomes had a significantly lower risk of antibiotic use. However, among Hispanic individuals and those of another race or ethnicity, greater income substantially increased risk. This may be because high income can afford greater capacity to negotiate local structural deficits (e.g., low provider density or transportation barriers), allowing personal resources to facilitate care-seeking. Indeed, this is supported by findings that greater income is associated with care-seeking for individuals with diarrhea [30]. Income did not impact antibiotic use in White individuals, but higher education reduced it, as it did in respondents of another race or ethnicity, consistent with a connection between education and health literacy. Insurance status was not associated with antibiotic use in any population, suggesting that income and education are stronger determinants of use. The qualitatively different socioeconomic associations we identified indicate a need to disentangle the population-specific effects of health literacy and resource availability.

Neighborhood characteristics also demonstrated substantial associations with antibiotic use in most populations. Greater social vulnerability (i.e., SVI) was associated with increased risk of antibiotic use among Hispanic respondents. Language barriers and logistical hurdles, which may be more common in communities with high SVI, have been noted as drivers of non-prescription antibiotic use in this population [31]. We cannot distinguish between prescribed and non-prescribed antibiotics in this study, though all those who used antibiotics had sought care. Examining specific community characteristics, we found that risk decreased by approximately 20% for each additional prescribing provider within 10km among Black individuals, which is consistent with prior research showing more antibiotic prescriptions, including inappropriate prescriptions, among Black children who lived in rural vs. urban settings [32]. Notably, neither rurality in that study nor lower provider density in our study were associated with greater antibiotic risk in the White population. Resource-constrained environments may suffer from a scarcity of specialists, which can lead to reliance on antibiotics [32], and our results would suggest that this particularly impacts Black populations. In contrast to this, we found that fewer personal vehicles in an area decreased the risk of antibiotic use among Black respondents, consistent with lack of transportation being a barrier to care-seeking [33]. It increased used among White respondents, suggesting access to public transit or other alternate transportation, such as in urban centers.

Age and sex are commonly linked to antibiotic use; however, the literature is mixed on their specific effects. Relative to children <5 years old, we found that children 5-17 years were at lower risk of antibiotic use among Black and other racial and ethnic groups, which is consistent with results across all races and ethnicities from a national study of antibiotic prescriptions for gastroenteritis [6] but opposite what studies for other conditions have found [20,34]. White adults had a lower risk of antibiotic use than children <5 years in our study, which is again consistent with results for all races and ethnicities in some [8] but not all [6] previous studies. Differences in parental expectations for antibiotics when seeking care for their children have been identified and could be driving the results we observed [35]. However, differences by child’s age, or comparison to the parent’s expectations for antibiotics when they seek care for themselves, have not been assessed. We identified no difference in antibiotic use between men and women, similar to previous work [6,8,15].

Some combination of clinical characteristics was important in each population: fever in Hispanic and White populations, immune status in Hispanic and other racial and ethnic groups, vomiting in Black and White individuals, and bloody diarrhea in other racial and ethnic groups. Immunocompromised individuals and those with fever more commonly experience severe disease [2], which may be prompting care-seeking and prescription. Vomiting is more frequently associated with viral illnesses and should thus reduce antibiotic prescriptions. These associations were in the expected directions. Bloody diarrhea is often associated with bacterial gastroenteritis and severe disease [2], but we found an inverse association with antibiotic use in one population. This could reflect its presence in noninfectious gastroenteritis, the contraindication to antibiotics among individuals with Shiga toxin-producing *Escherichia coli*, or a combination of factors [2]. As we had no data on pathogen testing results, we were unable to determine whether the differences in symptoms associated with antibiotic use were driven by underlying differences in gastroenteritis etiology. This could account for the differences we observed, but they may also have been driven by cultural differences in symptom perception [36] or statistically correlated symptoms.

Our use of the FoodNet Population Survey enabled us to conduct a nationally representative, population-based analysis; however, certain vulnerable populations may have been underrepresented. We used survey weights to remedy this to the extent possible; however, low cell counts necessitated aggregation of Asian, Pacific Islander, American Indian, Alaska Native, and multiracial populations. Studies able to disaggregate these populations are necessary to improve antimicrobial stewardship [9]. Additionally, the study could not account for all potential confounders, including detailed clinical histories that might justify antibiotic use. To focus attention on the greatest reasons for racial and ethnic disparities in antimicrobial use, we examined antibiotic use across the full illness process. This decision precluded us from directly comparing our estimates of antibiotic use to those obtained in specific medical settings (e.g., ambulatory visits [6]), because our analysis included individuals who did not seek care. It also precluded us from investigating factors such as care setting and provider characteristics [37,38]. These should be included in future studies, as implicit biases in clinical decision-making [39] and population-specific communication dynamics during medical encounters [40] can contribute to higher antibiotic prescribing.

## CONCLUSION

We identified substantial disparities in antibiotic use for gastroenteritis across major racial and ethnic groups in the U.S., placing these populations at greater risk of antibiotic-related adverse events. Understanding why a disparity exists is essential to remediating it. For each racial and ethnic group, we examined potential demographic, socioeconomic, clinical, and social environment characteristics associated with antibiotic use. We found that drivers of antibiotic use varied meaningfully by group, emphasizing the importance of disaggregating the causal factors of antibiotic use, as different populations will need different antimicrobial stewardship strategies [9], such as tailored clinical decision support tools and community-engaged education efforts.

## Data Availability

The data are restricted and cannot be shared by the authors. Access may be obtained from the CDC through application and a data use agreement. Analytic code (and derived, non-identifiable summary outputs) can be made available upon reasonable request to the corresponding author.

## ACKNOWLEDGEMENTS

We gratefully acknowledge coding assistance from Habibat Oguntade. We also thank the Foodborne Diseases Active Surveillance Network, Centers for Disease Control and Prevention, which supplied the data for this study. The findings and conclusions in this report are those of the authors and do not necessarily represent the official position of the Centers for Disease Control and Prevention. Data is available from CDC upon agreement, at their discretion.

## Author Contributions

Conceptualization: G.A.M.T. Methodology: D.K., F.R., G.A.M.T. Data curation: D.K., R.I., F.R. Formal analysis: D.K., R.I. Visualization: D.K. Writing – original draft: D.K. Writing – review and editing: R.I., F.R., G.A.M.T. Funding acquisition: F.R., G.A.M.T. Supervision: F.R., G.A.M.T. Project administration: F.R., G.A.M.T.

