## Supplemental Tables and Figures for "Racial and Ethnic Differences in the Determinants of Antibiotic Use for Acute Gastroenteritis in the United States"

*Supplement*

### FIGURES

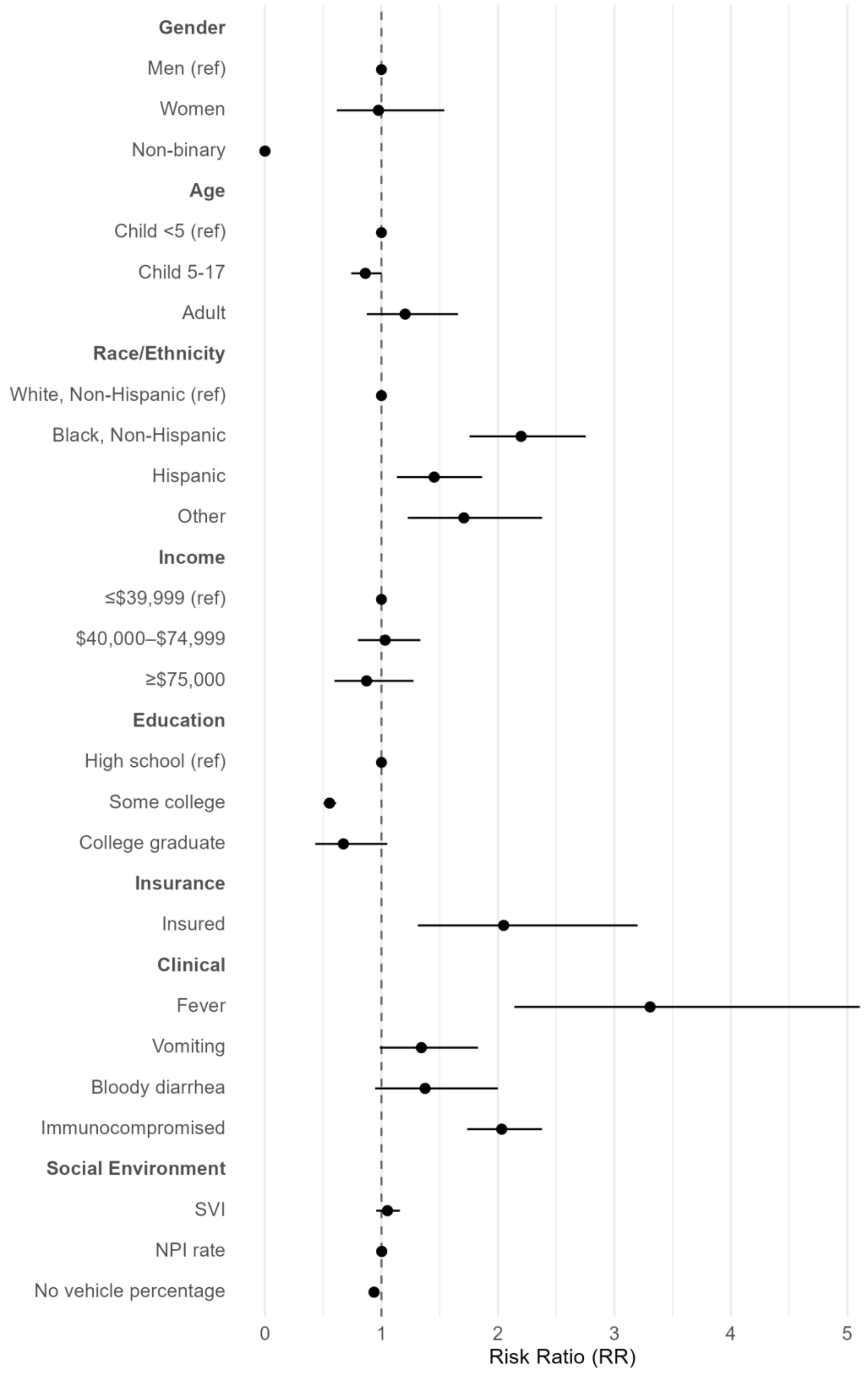
Gender
Men (ref)
Women
Non-binary
Age
Child <5 (ref)
Child 5-17
Adult
Race/Ethnicity
White, Non-Hispanic (ref)
Black, Non-Hispanic
Hispanic
Other
Income
≤$39,999 (ref)
$40,000–$74,999
≥$75,000
Education
High school (ref)
Some college
College graduate
Insurance
Insured
Clinical
Fever
Vomiting
Bloody diarrhea
Immunocompromised
Social Environment
SVI
NPI rate
No vehicle percentage
0
1
2
3
4
5
Risk Ratio (RR)

**Supplementary Figure 1. Risk ratios for antibiotic use of demographic, socioeconomic, clinical, and social environment characteristics among individuals reporting gastroenteritis in the past 7 or 30 days.** Risk of use was elevated for individuals identifying as non-Hispanic Black, Hispanic, or another race or ethnicity, relative to non-Hispanic White individuals. Dots indicate risk ratio (RR) point estimates and bars are 95% confidence intervals accounting for survey weights. For variables with >2 categories, the reference category is indicated, and for binary variables, the reference group is the absence of the characteristic. The RR for Social Vulnerability Index (SVI) is presented as the relative change in the proportion of individuals using antibiotics for each additional 0.1 increment in the index. The number of providers with a National Provider Identifier (NPI) per 1,000 population within a 10-km radius of the home zip code is presented as the relative change for each additional 1 provider. The percentage of a county with no personal vehicle is presented as the relative change for each additional 1%. Two versions of the survey were administered, one asking about health conditions in the previous 7 days and one about health conditions in the previous 30 days. Only the 7-day sample was used in the primary analysis due to greater potential of recall bias in the 30-day sample. We combined the samples in this sensitivity analysis.

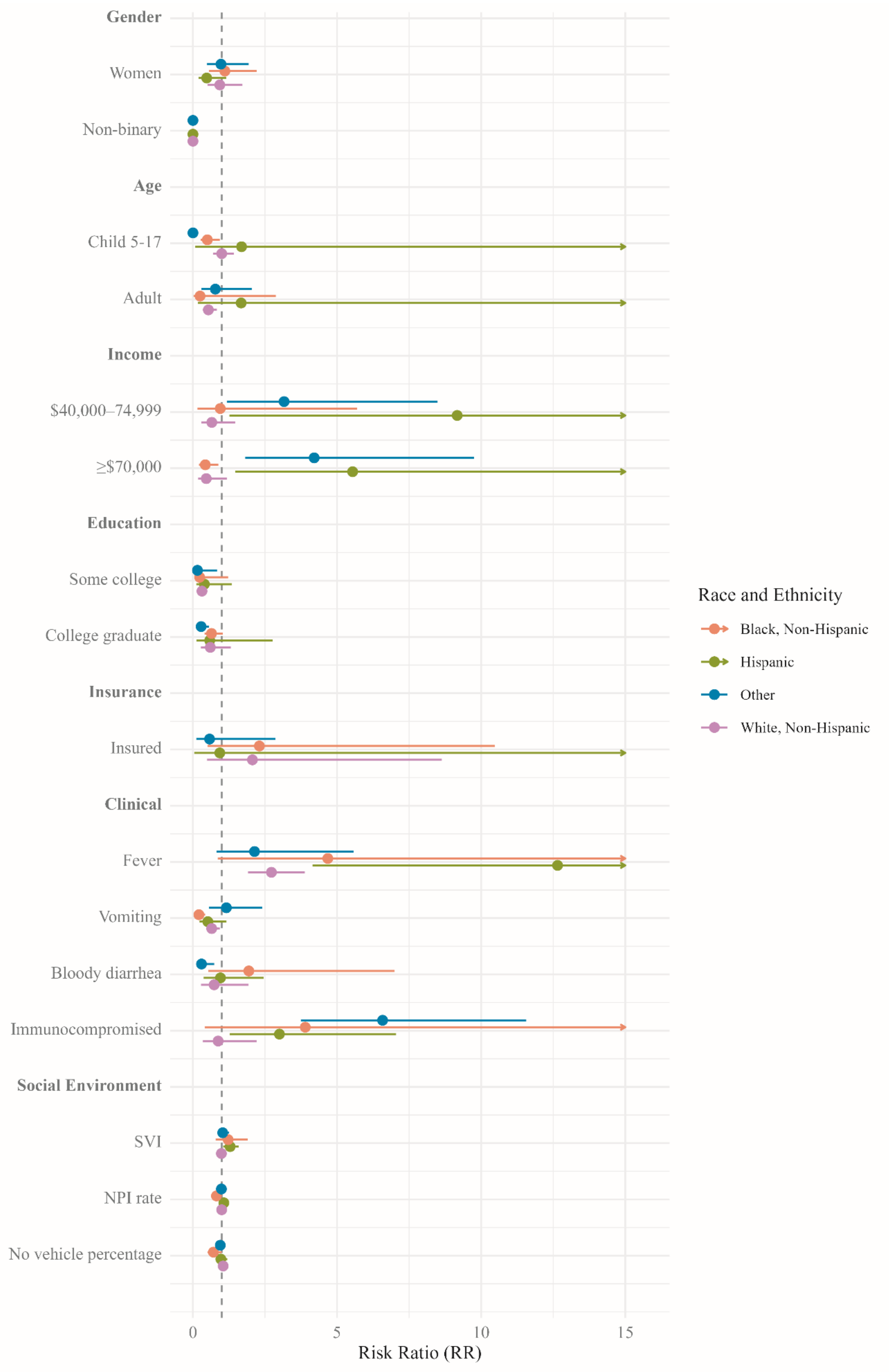

Gender
Women
Non-binary
Age
Child 5-17
Adult
Income
$40,000–74,999
≥$70,000
Education
Some college
College graduate
Insurance
Insured
Clinical
Fever
Vomiting
Bloody diarrhea
Immunocompromised
Social Environment
SVI
NPI rate
No vehicle percentage
0
5
10
15
Risk Ratio (RR)
Race and Ethnicity
Black, Non-Hispanic
Hispanic
Other
White, Non-Hispanic

**Supplementary Figure 2. Risk ratios for antibiotic use of demographic, socioeconomic, clinical, and social environment characteristics among individuals reporting gastroenteritis in the past 7 or 30 days, stratified by race and ethnicity.** Drives of antibiotic use for gastroenteritis varied by race and ethnicity. Dots indicate risk ratio (RR) point estimates and bars are 95% confidence intervals (CIs) accounting for survey weights. Arrows on 95% CIs indicate that the right-hand side of the CI exceeds 15. For variables with >2 categories, reference categories are shown in Figure 1, and for binary variables, the reference group is the absence of the characteristic. RRs for Social Vulnerability Index (SVI) are presented as the relative change in the proportion of individuals using antibiotics for each additional 0.1 increment in the index. The number of providers with a National Provider Identifier (NPI) per 1,000 population within a 10-km radius of the home zip code is presented as the relative change for each additional 1 provider. The percentage of a county with no personal vehicle is presented as the relative change for each additional 1%. Two versions of the survey were administered, one asking about health conditions in the previous 7 days and one about health conditions in the previous 30 days. Only the 7-day sample was used in the primary analysis due to greater potential of recall bias in the 30-day sample. We combined the samples in this sensitivity analysis.

### TABLE

**Supplementary Table 1.** Self-identified race of the 168 individuals classified as another race or ethnicity, 'Other'

| | **N (%)** |
|---|---|
| Black or African American | 1 (0.6) |
| American Indian or Alaska Native | 20 (11.9) |
| Asian | 56 (33.3) |
| Pacific Islander | 4 (2.4) |
| More than one race | 65 (38.7) |
| Other | 17 (10.1) |
| Unknown | 5 (3.0) |

**Supplementary Table 2.** Characteristics of participants who reported experiencing gastroenteritis in the previous 30 days, including individuals in the primary analysis who reported gastroenteritis in the previous 7 days, stratified by racial and ethnic group

| | Black, Non-Hispanic (n=272) | Hispanic (n=411) | Other (n=428) | White, Non-Hispanic (n=4,185) |
|---|---|---|---|---|
| **Antibiotic use, n (%)** | | | | |
| **No** | 241 (88.6) | 360 (87.6) | 393 (91.8) | 3,877 (92.6) |
| **Yes** | 31 (11.4) | 51 (12.4) | 35 (8.2) | 308 (7.4) |
| **Age, n (%)** | | | | |
| **Child <5** | 19 (7.0) | 34 (8.3) | 47 (11.0) | 195 (4.7) |
| **Child 5-17** | 26 (9.6) | 90 (21.9) | 83 (19.4) | 391 (9.3) |
| **Adult** | 227 (83.5) | 287 (69.8) | 298 (69.6) | 3,599 (86.0) |
| **Gender, n (%)** | | | | |
| **Man** | 86 (32.3) | 173 (43.5) | 172 (41.6) | 1,730 (42.1) |
| **Woman** | 180 (67.7) | 223 (56.0) | 237 (57.4) | 2,361 (57.5) |
| **Non-binary** | 0 (0.0) | 2 (0.5) | 4 (1.0) | 14 (0.3) |
| **Education, n (%)** | | | | |
| **High school** | 60 (22.1) | 91 (22.1) | 62 (14.5) | 681 (16.3) |
| **Some college** | 79 (29.0) | 124 (30.2) | 92(21.5) | 1,071 (25.6) |
| **College graduate** | 133 (48.9) | 196 (47.7) | 274 (64.0) | 2,433 (58.1) |
| **Income, n (%)** | | | | |
| **≤$39,999** | 107 (44.2) | 124 (33.5) | 105 (27.4) | 935 (25.6) |
| **$40,000-74,999** | 75 (31.0) | 99 (26.8) | 91 (23.8) | 943 (25.8) |
| **≥$75,000** | 60 (24.8) | 147 (39.7) | 187 (48.8) | 1,771 (48.5) |
| **Insurance, n (%)** | | | | |
| **No** | 24 (8.9) | 26 (6.4) | 18 (4.2) | 148 (3.6) |
| **Yes** | 247 (91.1) | 380 (93.6) | 407 (95.8) | 3,999 (96.4) |
| **Fever, n (%)** | | | | |
| **No** | 224 (88.2) | 310 (78.7) | 334 (81.9) | 3,438 (87.3) |
| **Yes** | 30 (11.8) | 84 (21.3) | 74 (18.1) | 501 (12.7) |
| **Vomiting, n (%)** | | | | |
| **No** | 167 (67.6) | 224 (61.9) | 260 (67.5) | 2,942 (76.6) |
| **Yes** | 80 (32.4) | 138 (38.1) | 125 (32.5) | 898 (23.4) |
| **Bloody diarrhea, n (%)** | | | | |
| **No** | 177 (92.2) | 261 (94.6) | 290 (95.7) | 3,206 (96.8) |
| **Yes** | 15 (7.8) | 15 (5.4) | 13 (4.3) | 105 (3.2) |
| **Immunocompromised, n (%)** | | | | |
| **No** | 240 (88.2) | 368 (89.5) | 385 (90.0) | 3,759 (89.8) |
| **Yes** | 32 (11.8) | 43 (10.5) | 43 (10.0) | 425 (10.2) |
| **SVI, mean (SD)** | 0.55 (0.22) | 0.56 (0.25) | 0.45 (0.21) | 0.43 (0.23) |
| **Providers within 10 km radius (per 1,000** | 8.42 (8.89) | 7.04 (6.43) | 8.19 (7.02) | 8.23 (8.21) |

| | | | | |
|---|---|---|---|---|
| **population), mean (SD)** | | | | |
| **Proportion of county population with no vehicle, mean (SD)** | 0.05 (0.04) | 0.03 (0.04) | 0.04 (0.05) | 0.03 (0.03) |
